# Agentic Artificial Intelligence as a Catalyst for Administrative Modernization: The Beginning of the End for Traditional Fax Workflows in Healthcare

**DOI:** 10.64898/2026.07.27.26359032

**Authors:** Anthony L. Lin, Sarah Curtis, Matthew Fitzsimmons, Nick Nguyen, Alexeis Baqui, Arch Desai, Lucy Stalker, Nathan Aycock, Srirama Koneru, Baskar Sridharan, Harry Phillips, Sreekanth Vemulapalli, Manesh R. Patel

## Abstract

**Background:** Healthcare has witnessed administrative staffing roles balloon to twice the number of employed clinicians, resulting in $950 billion per year in administrative costs to deliver healthcare. Administrative workflows, like fax routing, are ripe for automation given the high human labor cost necessary to complete these tasks. Fax transmissions remain a key mode of communication in modern healthcare, requiring substantial manpower, and incurring significant, though not well-characterized, costs to health systems. Opportunities may exist for agentic artificial intelligence (AI) to automate this administrative task.

**Methods:** This quality improvement study was performed in 2 phases at Duke University’s Division of Cardiology, a single tertiary-care cardiac referral center. The first retrospective phase employed an observational time study design surveying manual fax routing processes at 3 representative cardiology clinics from April 1, 2024, to July 12, 2024. The second phase quantified all inbound faxes received through the division’s communication hub from July 1, 2025, to December 31, 2025, and applied direct labor costs observed in the time study to quantify the economic burden of manual fax routing across the hub.

**Results:** The observational time study (Phase 1) demonstrated fax routing processing times ranging from 4.4 to 9.4 minutes depending on fax type, with a mean processing time of 6.0 minutes per fax. On average, the ambulatory clinics received 1,694 faxes per month and spent 169.1 person-hours routing faxes. The divisional communication hub (Phase 2) received 24,420 faxes over the study period, averaging 4,070 inbound faxes and 13,341 pages of information per month. Extrapolating direct labor efforts from the time study, 407 person-hours per month were spent processing inbound faxes. For our institution, this translated to $10,663.40 in total monthly costs, roughly 2.5 full-time equivalents.

**Conclusion:** Manual fax routing represents a substantial, measurable, and previously under-characterized operational and administrative burden. Given the significant opportunity to reduce labor time and costs, our study establishes fax routing as a high-value target for automation. Future work is needed to determine the impact of AI-automated fax routing on the time, labor, accuracy, and economics within clinical settings.

## INTRODUCTION

Total administrative spending in healthcare in the United States (US) is estimated to be $950 billion per year.^1^ While administrative overhead is necessary to run any organization, many US industries have managed to maintain a ratio of approximately 0.85 administrative workers for every employee in an industry-specialized role.^2^ Comparatively, US healthcare has witnessed administrative staff balloon to a sum twice as large as the number of employed physicians and nurses, incurring substantial labor costs associated with delivering healthcare.^2^

Artificial intelligence (AI), particularly agentic AI, with its ability to autonomously coordinate and execute multi-step processes without human prompting, has been an attractive tool to curb administrative costs in healthcare. While some estimates suggest these tools could generate savings as high as $200 to $360 billion in healthcare,^3^ these cost savings have yet to be fully realized. Additionally, alongside the growing shortage of healthcare workers nationally and globally,^4–6^ introducing autonomous AI tools to tackle repetitive and routine administrative tasks remains an attractive solution to help mitigate the consequences of these labor shortages. To better identify opportunities where agentic AI could improve administrative efficiency, realize cost savings, and liberate human knowledge workers, each administrative need requires thoughtful upfront quantification.

Facsimile (fax), a technology invented in the 1800s, remains an integral communication channel for healthcare. In fact, commercial estimates suggest that 70% of all healthcare communication still occurs via fax, resulting in over 9 billion pages of health information exchanged annually.^7^ The majority of faxes still involve human processing to manually collect and sort; match to the health record; and route to the appropriate destination. Not only does each step add labor time, the entire process is error-prone^8^ and associated with delays to patient care.^9,10^ Unfortunately, faxed documents remain a major source of administrative burden,^11^ serving as a significant driver of healthcare expenditure and staff burnout.

While AI automation of triage and routing workflows could meaningfully improve operational efficiency, the true operational burden of fax processing has not been well-characterized and it remains difficult to quantify the scale of opportunity and any associated returns on investment. We, therefore, sought to retrospectively characterize the administrative burden of fax routing for a communication hub at a large academic medical center prior to implementation of an AI automated fax routing workflow.

## METHODS

This was a retrospective quality improvement study performed in 2 phases at Duke University’s Division of Cardiology, a tertiary-care cardiac referral center. Phase 1 employed an observational time study design to survey the manual fax routing process at 3 representative cardiology clinics at Duke University from April 1, 2024, to July 12, 2024. The volume of inbound faxes, types of faxes received, and time associated with manual processing of each fax was recorded. Faxes were categorized into 1 of 7 fax categories based on the nature of the fax: Release of Information, Medication, Care Coordination, Clinical Documents, Administrative Forms, Monitor Notification, and Other (**Supplemental Table 1**). All time study data during this period were collected via direct observation.

Phase 2 was retrospectively performed on fax data from July 1, 2025, to December 31, 2025, and quantified all inbound faxes received through the division’s communication hub during the period. As the fax routing workflow did not change between the study’s two phases, average processing times observed in the original time study were applied to the total fax volume received by the communication hub to quantify total monthly and annual labor costs related to the manual fax routing workflow. The application of clinic-derived fax processing times to the communication hub was supported by use of a shared fax type taxonomy across both settings and comparable staff roles performing the routing workflow. Direct financial costs were estimated using the annual salary of a medical records specialist performing this work within a standard 40-hour workweek.

Retrospective categorization of fax type for all inbound faxes received during the study period was performed using a proprietary document processing pipeline developed by Trase Systems (www.trase.ai). The pipeline implemented a deterministic, multi-step workflow designed for healthcare document triage. For each inbound fax, the pipeline executed the following sequential steps: (1) text extraction from the fax document image using a vision language model; (2) generation of a structured document summary; (3) classification into 1 of 7 primary fax categories (Release of Information, Medication, Care Coordination, Clinical Documents, Administrative Forms, Monitor Notification, or Other) and corresponding subcategories; (4) extraction of protected health information elements for patient matching; and (5) condition-specific flagging (e.g., Left Ventricular Assist Device status, heart transplant status) via matched clinical codes. Each step utilized large language models operating within a fixed, protocol-driven workflow to ensure deterministic execution across all documents. The pipeline was run as an isolated instance separate from the production environment during off-peak hours to avoid interference with clinical operations. Only deidentified metadata and classification outputs were retained for analysis; no patient-identifiable information was included in the analytic dataset.

Descriptive statistics were collected for all fax documents received in both phases of the study. Continuous variables were presented using means and standard deviation (SD) and categorical variables were presented as frequencies. In Phase 1, the fax processing workflow for the 3 representative clinics was observed and measured to record human time spent processing each fax document, stratified by fax type. Primary outcomes of this phase included the number of occurrences of each fax type, processing time for each fax document, and total monthly time spent processing each fax type. In Phase 2, the total inbound fax volume for the communication hub was retrospectively categorized based on the fax taxonomy. Primary outcomes of this phase included the number of occurrences of each fax type. Given the shared fax taxonomy and workflow, the measured processing times from Phase 1 were applied to the fax documents received in Phase 2 to estimate the total monthly labor burden for processing all faxes received through the communication hub (**Figure 1**).

**Figure 1.**
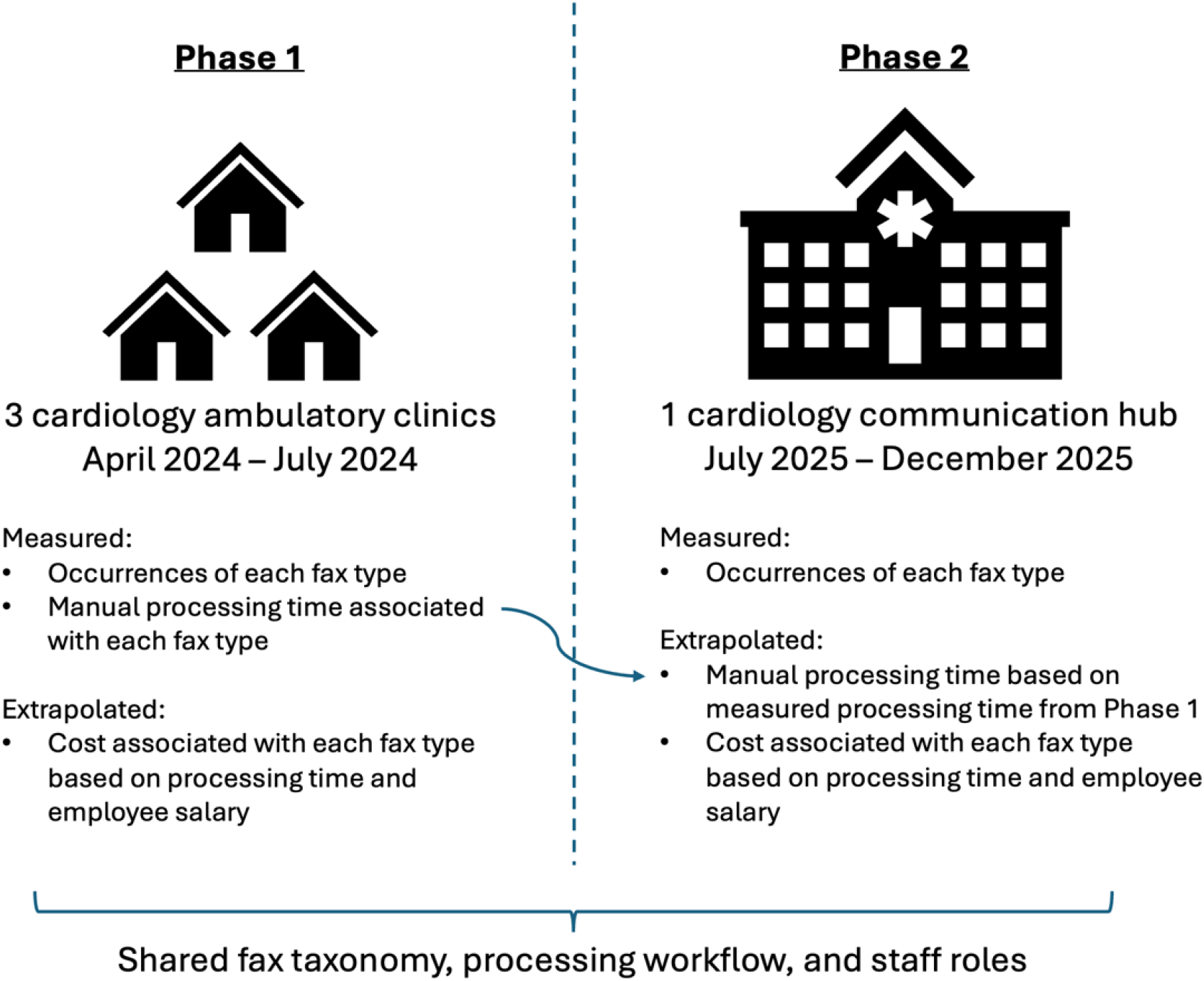
Diagram illustrating the measured and extrapolated outcomes from Phase 1 and Phase 2. Given the shared fax taxonomy and workflow, measured manual processing times in Phase 1 were applied to the fax documents received in Phase 2 to estimate total labor burden.

All statistical analyses were performed using R 4.5.1 (R Core Team, 2025). As this was a quality improvement study without direct access to protected health information, the study was exempt from Institutional Review Board (IRB Pro00120244). The Strengthening the Reporting of Observational studies in Epidemiology (STROBE) guidelines were followed.^12^

## RESULTS

### Phase 1: Observational Time Study of Representative Cardiology Clinics

The observational time study was conducted inside 3 ambulatory clinics within the Division of Cardiology to better understand the processing time required to complete the fax routing workflow. These clinics support 79 providers (physicians and advanced practice providers) throughout the week, performing approximately 200 patient clinic appointments per day. All their faxes were initially received and digitized into a central fax inbox. Next, medical records specialists manually reconciled each fax with the electronic health record to identify the correct patient and provider. Employees then reviewed the fax’s content, consulted clinic protocols, and routed that fax to the appropriate clinical team (**Figure 2**).

**Figure 2.**
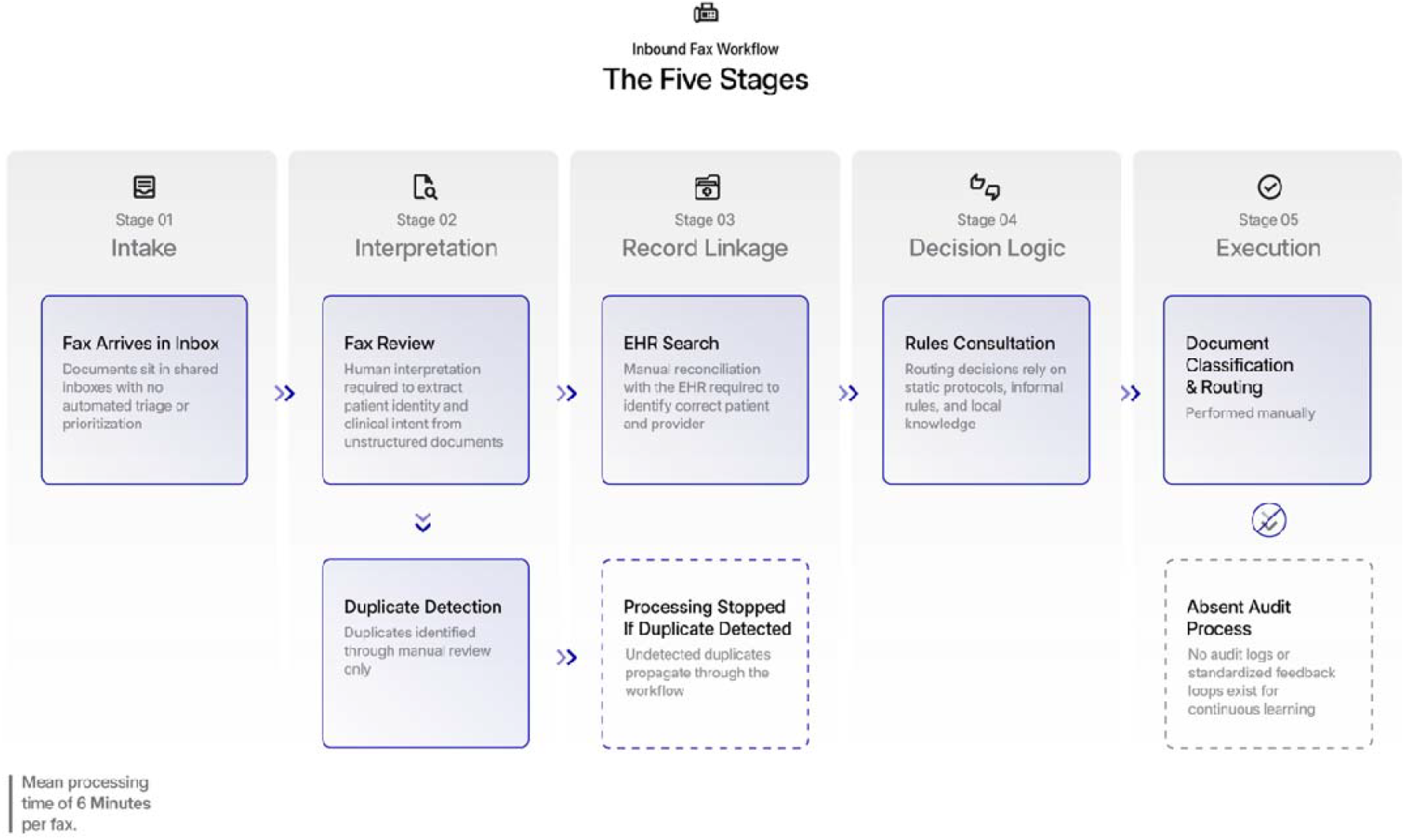
Flow diagram of the fax routing workflow performed by human employees. Mean processing time was measured to be 6.0 minutes per fax document. EHR = electronic health record.

On average, 1,694 faxes were received by the clinics monthly, each with a mean processing time per fax of 6.0 minutes to perform the entire routing workflow (**Table 1**). Medication-related faxes required the least amount of time to process (4.4 minutes) while Clinical Documents-related faxes required the most amount of time to process (9.4 minutes). Total time consumed by manual fax routing for the clinics was 169.1 person-hours monthly, equivalent to approximately 1 full-time equivalent (FTE) of work. At a median medical records specialist salary of $50,250 annually,^13^ this direct labor cost translated to $4,425.66 per month to perform the task of fax routing at this single clinic.

**Table 1.** Processing time by fax type at a single clinic based on observational time study performed April – June 2024. *Monitor Notification faxes were not fully integrated into the fax workflow in 2024 so no data were presented for this fax type.

|  | Number of<br>faxes per<br>month | Mean<br>processing time<br>per fax, minutes | Total monthly<br>processing time,<br>hours |
| --- | --- | --- | --- |
| Fax Type |  |  |  |
| Release of Information | 57 | 6.0 | 5.7 |
| Medication | 834 | 4.4 | 61.5 |
| Care Coordination | 270 | 6.0 | 27.0 |
| Clinical Documents | 329 | 9.4 | 51.3 |
| Administrative Forms | 159 | 7.0 | 18.5 |
| Monitor Notification | * | * | * |
| Other | 47 | 6.7 | 5.2 |
| **Total per month** | **1,694** | **6.0** | **169.1** |

### Phase 2: Quantification of Total Inbound Fax Volume from Communication Hub

In total, the division’s communication hub received 24,420 faxes over the last 6 months of 2025, averaging approximately 4,070 inbound faxes per month with an SD of 1,892 faxes (**Table 2**). The highest volume of faxes received were Medication-related (averaging 2,976 faxes per month) while Care Coordination-related faxes were the longest (average 14.5 pages per fax). The highest volume senders of faxes were pharmacies (61%), clinics and hospitals (21%), and insurers (13%). On average, each inbound fax contained 3.3 pages (inclusive of the cover page) with an SD of 8.1 pages, resulting in a mean of 13,431 pages faxed monthly for the single communication hub. Most (54%) inbound faxes contained only 1 page (n=13,141). However, 13% of faxes contained ≥5 pages, with one fax containing 317 pages.

**Table 2.** Total volume of inbound faxes received within the division’s communication hub.

| **Fax Type** | **Faxes per month (mean ± SD)** | **Pages per fax (mean ± SD)** |
| --- | --- | --- |
| Release of Information | 190 ± 104 | 3.6 ± 2.8 |
| Medication | 2,976 ± 1,364 | 1.6 ± 1.8 |
| Care Coordination | 454 ± 59 | 14.5 ± 19.4 |
| Clinical Documents | 280 ± 148 | 6.9 ± 12.9 |
| Administrative Forms | 194 ± 100 | 2.9 ± 2.1 |
| Monitor Notifications | 7 ± 5 | 4.6 ± 8.2 |
| Other | 47 ± 24 | 2.0 ± 3.0 |
| **Total** | **4,070 ± 1,892** | **3.3 ± 7.9** |

Modelling of associated labor costs was performed by applying average processing times experienced during the observational time study for each fax type. At a median medical records specialist salary of $50,250 annually,^13^ the direct labor costs ranged from $1.92-$4.10 per fax processed, depending on fax type. The total monthly direct labor costs for fax routing at our division’s communication hub was approximately $10,663.40 per month (**Table 3**).

**Table 3.** Economic analysis of direct labor costs for fax routing at one division’s communication hub. *Monitor Notification faxes were not fully integrated into the fax workflow in 2024 for time study data so no economic estimates were presented for this fax type. †Mean processing time per fax extrapolated from observational time study data performed from April – June 2024 (Table 1).

| **Fax Type** | **Mean processing time per fax, minutes†** | **Total monthly processing time, hours (mean ± SD)** | **Mean labor cost per fax, \$** | **Total monthly labor cost, \$ (mean ± SD)** |
| --- | --- | --- | --- | --- |
| Release of Information | 6.0 | 19.0 ± 10.4 | 2.62 | 497.80 ± 272.48 |
| Medication | 4.4 | 218.2 ± 100.0 | 1.92 | 5,713.92 ± 2,618.88 |
| Care Coordination | 6.0 | 45.4 ± 5.9 | 2.62 | 1,189.48 ± 154.58 |
| Clinical Documents | 9.4 | 43.9 ± 23.2 | 4.10 | 1,148.00± 606.80 |
| Administrative Forms | 7.0 | 22.6 ± 11.7 | 3.05 | 591.70 ± 305.00 |
| Monitor Notifications | * | * | * | * |
| Other | 6.7 | 5.2 ± 2.7 | 2.92 | 137.24 ± 70.08 |
| **Total** | 6.0 | 407.0 ± 189.2 | 2.62 | 10,663.40 ± 4,957.04 |

## DISCUSSION

In summary, our study estimated that manual fax routing consumed 4,884 person-hours annually for Duke University’s Division of Cardiology communication hub, the equivalent of 2.5 dedicated FTEs. This resulted in approximately $128,000 in annual direct labor costs to perform manual fax routing at our division’s communication hub alone.

Although the calculations may seem unique to one process, the fax routing use case in this study demonstrates a workflow amenable to automation. Fax routing management is an example pertinent to healthcare entities (pharmacies, clinical offices, hospitals, rehabilitation facilities, prior authorization, procedural clearance, etc.) experiencing redundancies, loss time, inefficiencies, and delayed care caused by fragmented communication links.

Given the fax burden observed for Duke University’s Division of Cardiology, which supports 140 full-time physicians, when scaled to the 2,800 full-time physicians across Duke University Health System, we estimate our system-wide fax burden to total 976,800 faxes per year, requiring 97,680 person-hours and approximately $2.5 million in human labor costs to manually sort, match, and route faxes. With an estimated 412 academic medical centers and hospitals^14^ experiencing a fax burden similar to ours, this amounts to 402 million faxes and 1.3 billion pages of information consuming 40 million person-hours nationally to facilitate healthcare communication throughout the US.

While AI holds significant promise for reducing administrative burden, its deployment in revenue-oriented workflows to date has sparked an adversarial “arms race” between payors and providers.^15^ While ambient AI scribes have supported higher-complexity medical coding,^16^ payors have responded by automatically downcoding level 4 and 5 Evaluation/Management codes unless documentation explicitly supports higher complexity.^17,18^ While payors have used AI to assist with streamlining insurance denials,^19^ providers and health systems are increasingly turning to the same tools to help appeal these decisions.^20^

Applying AI to purely operational and logistical administrative workflows, such as automated document classification and fax routing, delivers direct cost savings by reducing manual labor costs without triggering an adversarial response. In fact, one implementation study at a large academic medical center recently demonstrated the technical feasibility of an AI-enabled document classifier for managing fax workflows, estimating an 80-90% reduction in cost to perform the task of fax routing compared to competitor pricing for similar work.^21^ Another academic medical center recently implemented an AI solution to triage faxed referrals for urgency and managed to reduce its turnaround time for urgent referrals from 33 hours to 1 hour.^22^ While many complexities and challenges hindering the operationalization of agentic AI in healthcare administration remain,^23^ an agentic AI routing solution tied to other tools holds potential to start the liberation process from costly and repetitive administrative tasks within healthcare settings.

This study has limitations worth discussing. First, this was a study of administrative fax burden on a single division’s communication hub within a single academic medical center. While our cardiology division may be representative of administrative volumes seen at other large tertiary-care academic medical centers, it may not generalize to other institutions with different workflows and referral patterns. Second, our estimations of labor costs do not capture cases in which fax routing required rework, further review, correction of errors, or downstream costs from delays. As such, the labor costs presented in this study are conservative estimates and the true costs experienced by institutions may be higher. Additionally, all administrative overhead and indirect costs were excluded from the analysis, which further contribute to costs incurred by an institution. Finally, this was a retrospective study on fax volumes received with estimated processing times based on representative time study data from 3 ambulatory clinics. While the time study provides representative processing times that apply to the full volume of faxes received through the communication hub, it does not account for workflows or processes that may have changed within the communication hub that may impact labor costs.

In conclusion, manual fax routing represents a substantial, measurable, and previously under-characterized operational and administrative burden. Given the significant opportunity to improve labor time and capital cost, fax routing is one administrative challenge that may be ripe for AI automation in healthcare. Furthermore, automated fax routing may enable reallocation of clinician effort from administrative tasks to higher-value activities, including clinical decision-making, patient interactions, and care coordination—an enduring objective of healthcare delivery that has thus far remained difficult to operationalize at scale.

## Data Availability

All data produced in the present work are contained in the manuscript

