## Supplementary Materials for "Agentic Artificial Intelligence as a Catalyst for Administrative Modernization: The Beginning of the End for Traditional Fax Workflows in Healthcare"

| **Fax Type** | **Subcategories** |
| --- | --- |
| Release of Information | Release of Information, Prior Authorization, Approved Authorizations, Catheterization Pending, Catheterization Denials |
| Medication | Prescription Refill, Medication Clarification, Medication Non-Adherence Advisory, Authorization to Stop Medication, Medication Recommendation, Medication Prior Authorization, Medication Contraindication |
| Care Coordination | Cardiology Referral, Cath Referral, Coronary CTA Referral, Cardiac Consults, Home Health Request, Home Health, Heart Transplant Coordination, Left Ventricular Assist Device Coordination |
| Clinical Documents | Discharge Summary, Progress Notes, Critical Results, Imaging Reports, Lab Results (External), Electrocardiograms |
| Administrative Forms | Disability Forms, Cardiac Rehabilitation Order Forms, Dental Clearance, Medical Clearance |
| Monitor Notification | Shipment, No Transmission, Enrollment, Notification |
| Other | Miscellaneous |

Supplemental Table 1. The taxonomy used for the study, listing each primary fax type and corresponding subcategories.
